# Early Repolarization Pattern and Suicidal Risks: A Single Center Case-Control Study

**DOI:** 10.1101/2020.07.10.20150482

**Authors:** Hiroshi Kameyama, Kenichi Sugimoto, Kyoko Itoh, Kazutaka Nukariya, Tomohiro Kato, Masahiro Shigeta

## Abstract

**Background:** Early repolarization pattern was reported to be associated with mental illness. However, its role in patients with the suicidal risks remains unclear.

**Objective:** The aim of this study is to examine the frequency of Early repolarization pattern in patients with the suicidal risks compared with the age matched medical checkups.

**Method:** A retrospective analysis of 27 patients with a history of suicide risks, including suicide attempt and nonsuicidal self-injury, and a family history of suicide. The presence of early repolarization pattern was compared in patients with suicidal risks and controls. Social and psychological factors were also compared among the patients with suicidal risks with or without Early repolarization pattern.

**Result:** Comparing the controls, Early repolarization pattern was significantly observed in the patients (14 patients: 52%, 7 controls: 9%, P<0.001). After logistic regression including the other clinical findings among the patients and controls, the presence of Early repolarization pattern was associated with the patients with the suicidal risks (p < 0.001). In the patients with suicide, there were no difference between the clinical factors compared with or without Early repolarization pattern.

**Conclusion:** There could be association between Early repolarization pattern and the patients with the suicidal risks. Further studies are needed to confirm the association between suicidal risks and Early repolarization pattern.

## Introduction

Suicide is a major public health concern and is among one of the leading causes of death. [1] In order to address one of today’s most challenging public health issues, it is important to find ways to predict and possibly prevent suicide.

Mental illness has been reported as the largest risk factor for suicide attempts.[2] However, suicide does not necessarily occur only with serious mental illness.[3] Although, several approaches having been made from various social, psychological, and psychiatric factors to prevent from completing suicide by intervening in people with the backgrounds, it is very difficult to predict suicide attempts even considering the associated factors in clinical practice.[4] The risk factors for suicide are under debate, however, it is clear that the history of suicide attempt itself is a risk of completion of suicide[5] and that Non-suicidal self-injury(NSSI) is related to suicide attempt[6], and the fact that family history of suicide is also considered to be risk of suicide attempt[7] suggests that suicide attempt has some genetic and biological basis.

In recent years interventions from a biological perspective have also been reported. For example, the patients with suicide attempt history and suicidal ideation showed high-gamma rhythm compared to the control group on Quantitative EEG.[8] On fMRI, a decrease in reactivity in the insular cortex could be the risk factor for suicide.[9] However, no solid conclusion could be made. Regarding these biological methods, there are also problems of time and cost, which are problems for practical application.

Recently, there have been reports on psychiatric disorders and Early repolarization pattern (ERP). It has been reported associations between psychiatric disorders such as Attention-deficit hyperactivity disorder (ADHD) [10, 11], serious mental illness and ERP.[12] These reports suggest the associations between ERP and psychiatric disorders. As, ERP and psychiatric disorders are associated, ERP may be observed more in patients with suicidal risks compared to controls without suicidal risks.

As mentioned above, suicide and mental illness are known to be strongly related, but no ECG studies have been conducted in patients with suicidal risks. This study was targeted at psychiatric patients, particularly those at high risk, who had a history of attempted suicide, NSSI. Additionally, we also included the patients with a family history of suicide considering biological aspects.

We hypothesized that ERP was significantly observed in patients with the suicidal risks in our hospital. The patients with a history of suicide attempts, patients who had history of NSSI, family history of suicide attempts within their second-degree relatives were investigated as the patients with the suicidal risks. The aim of this study is to compare the presence of ERP in the patients with the suicidal risks and medical checkups without mental illness or psychiatric family histories.

We also assessed the clinical factors including the social background, such as working, presence of cohabitant, marital status, with or without ERP.

## Methods

### Participants

We retrospectively investigated adult 27 patients in the Jikei Kashiwa hospital from January 2015 to July 2018 having the suicidal risks (History of a suicide attempt: 20 patients, NSSI:4 patients, family history of suicide within second degree: 3 patients.) without use of antidepressants, antipsychotics, or mood stabilizers. For the control groups, age matched 74 cases without psychiatric disorder or familial history within second degree relatives, who were randomly selected in medical checkup from Center for Predicitive Medicine of the Jikei university.

We excluded the electrocardiograms in hypothermia or hyperthermia, after post-cardiopulmonary resuscitation following suicide attempt, performed after overdose or poisoning suicide. The patients or the controls with organic heart disease, bundle branch block, and WPW syndrome [13], which could affect ERP. This study has been implemented with the approval of our ethics committee (approved No. 30-152).

### ECG Evaluation

All ECGs were recorded at a paper speed of 25 mm/sec and amplitude 1 mV/10 mm. Two independent reviewers (H.K: Medical Doctor and K.S: Cardiologist) blinded to clinical findings analyzed all the first recorded ECG without antipsychotic, mood stabilizer or antidepressant in the patient’s group and the first recorded ECG in the controls.In the evaluation of ERP, discrepancies were resolved by consensus. To classify an ECG as ERP, both reviewers had to be in agreement. For evaluating ERP, we followed the evaluation criteria proposed in 2015 by Peter W. Macfarlane as follows.[14] ERP is present if all the following criteria are met. First, there is an end-QRS notch or slur on the downslope of a prominent R-wave. If there is a notch, it should lie entirely above the baseline. The onset of a slur must also be above the baseline. Second, the J-point peak is ≥0.1 mV in two or more contiguous leads of the 12-lead ECG, excluding leads V1 to V3. Third, QRS duration is <120 ms. Both slur and notch should occur in the final 50% of the R-wave downslope to be regarded as ERP in order to exclude fragmented QRS.

### Confounding factors and Social and Psychological factors

We assessed age, sex, Heart rate, and coronary risk factors such as diabetes, hypertension, dyslipidemia, and smoking history. Moreover, among the patients, we also evaluated age, sex, heart rate and, social and psychological factors such as employment, the presence of cohabitant, marital status, with or without ERP.

### Power analysis

Power analysis, with β=0.20 and α=0.05, was conducted assuming a difference of 20% based on the previous studies [10,11]. We had a projected sample size of approximately 43 patients (total of 172 subjects).

### Statistical evaluation

We used IBM SPSS 25 for statistical analysis. The normality of the continuous variables was assessed using the Kolmogorov-Smirnov test. The continuous variables were compared using an unpaired Student’s T-test or the Mann–Whitney U-test when appropriate. Continuous data are expressed as mean ± SD for normally distributed variables or median [25th, 75th percentiles] for non-normally distributed variables. The categorical variables were compared using the chi-square analysis or fisher test when appropriate. Logistic regression analysis was performed to identify independent risk for suicidal risk among the patients and control by setting heart rate, sex, age, PR interval, QTc duration, and ERP as independent variables, and presence of the suicidal risks as the dependent variable. All tests were two-tailed, P value < 0.05 was considered to be statistically significant.

## Results

### Study participants and clinical characteristics

Table 1 shows that clinical characteristics of study participants. Regarding sex, diabetes, hypertension, dyslipidemia, and smoking history, there was no significant difference between the patients and the control groups. ECG findings showed that faster heart rate (74 ± 15 msec versus 64 ± 9 msec : P <0.001)and shorter PR interval(146 ± 20 msec versus 160 ± 32 msec: P=0.005) in the patients compared with in the medical checkups. In the patients, the QTc interval was significantly longer than the controls (423 ± 23 msec versus 411 ± 16 msec: P = 0.004). ERP was observed to be higher in the patients’ group than in the control group (14 versus 7: P <0,001). The patients had a trend toward higher amplitude of J peak (2.0 ± 0.6mm versus 1.5 ± 0.4 mm: P = 0.053). ERP was observed in the inferior leads and both inferior and lateral leads in the patients’ group No sudden cardiac death occurred during follow up.

**Table 1.** Comparison of clinical characteristic in the patients and controls.

|  | The Patients (N =27) | Controls (N = 74 ) | P Value |
| --- | --- | --- | --- |
| Females (%) | 18 (67%) | 33 (45%) | 0.071 |
| Age (years) | 45 ± 20 | 45 ± 21 | 0.997 |
| Coronary risk factors |  |  |  |
| Diabetes mellitus (%) | 1 (4%) | 1 (1%) | 0.465 |
| Hypertension (%) | 3 (11%) | 3 (4%) | 0.338 |
| Dyslipidemia (%) | 4 (15%) | 20 (27%) | 0.292 |
| Smoker (%) | 3 (11%) | 6 (8%) | 0.698 |
| ECG findings |  |  |  |
| Heart rate (bpm) | 74 ± 15 | 64 ± 9 | <0.001 |
| PR intervals (ms) | 146 ± 20 | 160 ± 32 | 0.005 |
| QRS duration (ms) | 92 [ 88-103 ] | 100 ± 14 | 0.072 |
| QTc intervals (ms) | 423 ± 23 | 411 ± 16 | 0.004 |
| ERP (%) | 14 (52%) | 7 (9%) | <0.001 |
| Amplitude of J peak (mm) | 2.0 ± 0.6 | 1.5 ± 0.4 | 0.053 |
| Inferior leads(%) | 7 (26%) | 6 (8%) | 0.038 |
| Left precordial leads(%) | 0 (0%) | 1 (1%) | 1.000 |
| Lateral leads(%) | 2 (7%) | 0 (0%) | 0.07 |
| Both inferior and lateral leads (%) | 5 (19%) | 0 (0%) | 0.001 |

ERP: Early repolarization pattern. Value as mean ± SD, no.(%) or median [25th, 75th percentiles].

Logistic regression analysis was performed to identify independent predictors for suicidal risk among the patients and controls by setting independent variable as the presence of the suicidal risks and dependent variables as sex, age, heart rate, PR interval, QTc duration, the presence of ERP. ERP and faster heart rate were also associated with the patients with suicidal risks excluding the other factors (ERP: 95 % confidence interval, 4.2–67.1, p < 0.001, Heart rate: 95% confidence interval, 1.01-1.12, P = 0.021) (table 2).

**Table 2.** Logistic regression among the patients to predict the presence of the suicidal risks.

| Variables | B coefficient | standard error | 95%CI of OR |  | *P* value |
| --- | --- | --- | --- | --- | --- |
|  |  |  | Lower bound | Upper bound |  |
| sex (female) | 0.484 | 0.681 | 0.427 | 6.162 | 0.477 |
| Age (year) | 0.005 | 0.015 | 0.975 | 1.035 | 0.76 |
| Heart Rate (per bpm) | 0.063 | 0.027 | 1.010 | 1.124 | 0.021 |
| PR interval (msec) | -0.026 | 0.016 | 0.944 | 1.006 | 0.107 |
| QTc (msec) | 0.009 | 0.016 | 0.979 | 1.041 | 0.554 |
| ERP (presence) | 2.822 | 6.391 | 4.211 | 67.100 | <0.001 |

CI: confidence interval, ERP: early repolarization pattern.

There were no differences in clinical findings including age sex heart rate and, the psychiatric background: suicide attempt, NSSI, family history of suicide, and social background, such as cohabitant, marital status, and employment with or without ERP in our study (Table3).

**Table 3.** Comparison of clinical characterics patients with or without ERP.

|  | Patients with ERP (N = 14) | Patients without ERP (N = 13) | P Value |
| --- | --- | --- | --- |
| Age (years) | 43 ± 22 | 47 ± 19 | 0.676 |
| Sex(female) | 7 (50%) | 11 (85%) | 0.103 |
| Heart rate (bpm) | 70 ± 13 | 78 ± 17 | 0.202 |
| Clinical characteristics |  |  |  |
| Suicide attempt | 10 (71%) | 10 (77%) | 1.000 |
| NSSI | 2 (14%) | 2 (15%) | 1.000 |
| Family history of suicide | 1 (7%) | 2 (15%) | 0.596 |
| Cohabitant (present) | 11 (78%) | 11 (84%) | 1.000 |
| Marriage (present) | 8 (30%) | 10 (77%) | 0.695 |
| Employed | 6 (43%) | 8 (62%) | 1.000 |

ERP:early repolarization pattern, NSSI: Non-suicidal self injury. Value as mean ± SD, no.(%) or median [25th, 75th percentiles]

## Discussion

In this study, ERP was found to be significantly higher in the group with suicidal risk than in the healthy group (P < 0.001). Heart rate was faster (P <0.001), PR interval was shorter(P=0.005) and QTc interval was longer(P=0.004) in the patients. ERP and faster heart rate were also associated with the patients with suicidal risks after logistic regression(P<0.001). There were no differences in the social or psychological background compared with or without ERP in this study.

### For these results we considered three possibilities

As a First consideration, we must consider the involvement of the autonomic nerves because the patients showed faster Heart rate, and shorter PR interval. It is known that vagal tone is reduced in patients with suicide ideation [15]. In contrast, Early repolarization pattern is associated with higher vagal tone [16]. However, interventricular conduction delays could masquerade as ERP due to reduced vagal tone, even if heart rate is in the normal range.

### Secondly, it may be explained in terms of ion channel abnormalities

In recent years, ion channel abnormalities have been pointed out in mental illness. Mental illness is strongly related to suicide as mentioned, it would be reasonable to think that suicide overlaps the biological basis of mental illness. CACNA1C gene mutations are associated with depression, bipolar disorder, and schizophrenia. [17-20] The relationship are reported between ABCC9 and familial delusional disorder.[21] These genes are known as genes related to cardiac ion channel diseases [22, 23] such as Early repolarization syndrome.

Considering these past reports there may be a genetic overlap between genetic mutations that can be a risk of suicide attempts and cardiac ion channel disease.

As a third possibility, chronic inflammation is also considered as the background of ECG abnormalities. Chronic inflammation is also known to be associated with suicide attempt history. Miná VA et al. reported that blood levels of IL-6 and TNF-α are elevated in depressed patients with strong suicidal ideation.[24] In autopsy after suicide attempts, microglial activity was seen in histological studies. [25] There are also reports on the relationship between chronic inflammation and electrocardiogram abnormalities. Stumpf et.al reported that the cases with ERP showed increased IL-6 in blood.[26] Additionally, Lazzerini PE et.al reported that autoantibodies block human Ether-a-go-go Related Gene(HERG) channel prolongation by blocking HERG channels.[27] In view of these facts, it is considered that chronic inflammation is associated as a background of ECG abnormalities in patients with a history of suicide attempts.

## Limitaiton

This study is a retrospective study in a single center. These results cannot be immediately applied for routine clinical use because of the retrospective nature of the study design and small sample size. Psychiatric patients without suicidal risks were not studied, which limits generalizability of the results. The genetic tests associating cardiac ion channels or evaluate for inflammatory markers including antibodies, CRP or IL-6 were not performed. We did not perform Heart Rate Variability to evaluate vagal tone. Since there are many cases in which multiple ECGs have not been performed in the control group and the patients, the effects of day-to-day variability and daily fluctuation could not be evaluated.

## Conclusion

This case-control study in a small population suggests that there may be associations between ERP and the suicidal risks. Further, studies considering for diagnosis of mental disorder, autonomic tones and genetic factors should be developed in the future.

## Data Availability

N/A

## Acknowledgments

None.

